# Infection-competent monkeypox virus contamination identified in domestic settings following an imported case of monkeypox into the UK

**DOI:** 10.1101/2022.06.27.22276202

**Authors:** Barry Atkinson, Christopher Burton, Thomas Pottage, Katy-Anne Thompson, Didier Ngabo, Ant Crook, James Pitman, Sian Summers, Kuiama Lewandowski, Jenna Furneaux, Katherine Davies, Timothy Brooks, Allan M Bennett, Kevin S. Richards

**Affiliations:** Research and Evaluation, UK Health Security Agency, Porton Down, Salisbury, United Kingdom; High Containment Microbiology, UK Health Security Agency, Porton Down, Salisbury, United Kingdom; Rare and Imported Pathogens Laboratory, UK Health Security Agency, Porton Down, Salisbury, United Kingdom; National Institute for Health Research, Health Protection Research Unit in Emerging and Zoonotic Infections, Liverpool, United Kingdom; Oxford Brookes University, Headington Campus, Oxford, United Kingdom

**Keywords:** Monkeypox, Monkeypox virus, Sampling Studies, Personal Protective Equipment, Communicable Diseases, Imported, Communicable Diseases, Emerging

## Abstract

An imported case of monkeypox was diagnosed in December 2019 in a traveller returning from Nigeria to the UK. Subsequently, environmental sampling was performed at two adjoining single room residences occupied by the patient and their sibling. Monkeypox virus DNA was identified in multiple locations throughout both properties, and monkeypox virus was isolated from several samples three days after the patient was last in these locations. Positive samples were identified following use of both vacuum and surface sampling techniques; these methodologies allowed for environmental analysis of potentially contaminated porous and non-porous surfaces via real-time quantitative PCR analysis in addition to viral isolation to confirm the presence of infection-competent virus.

This report confirms the potential for infection-competent monkeypox virus to be recovered in environmental settings associated with known positive cases and the necessity for rapid environmental assessment to reduce potential exposure to close contacts and the general public. The methods adopted in this investigation may be used for future confirmed cases of monkeypox in order to establish levels of contamination, confirm the presence of infection-competent material, and to identify locations requiring additional cleaning.

**Originality-Significance Statement:** Several imported cases of human monkeypox infection, an emerging infectious disease with a case fatality rate of up to 10%, have been identified in recent years including importations into the United Kingdom, the United States, Israel, and Singapore. It is likely that this phenomenon relates to decreased immunity against monkeypox infection in endemic regions that was previously provided via the smallpox vaccination programme. It is therefore likely that further imported cases of monkeypox will be reported in future; such occurrences will require significant clinical oversight, including suitable infection control measures. Environmental sampling to identify contaminated sites that may pose a risk can inform infection control guidance. This report documents an environmental sampling response following an imported case detected in late 2019 in the UK. Monkeypox virus DNA was readily identified in numerous locations throughout two domestic residences associated with the infected patient and infectious virus was isolated from several environmental samples confirming that contaminated environmental settings may pose a risk for onward transmission. The methods utilised in this report may advise future environmental responses following cases of this high consequence emerging viral disease in order to prevent secondary cases in close contacts or members of the public.

## Introduction

Monkeypox is a re-emerging zoonotic disease resulting from infection by monkeypox virus (MPXV), a double-stranded DNA virus classified within the *Orthopoxvirus* genus in the family *Poxviridae*. The clinical disease resulting from infection with MPXV is similar to smallpox but with marked lymphadenopathy; however, the symptoms and severity are typically far milder. Following an incubation period of approximately 5-14 days, early symptoms include fever, fatigue, myalgia, swollen lymph nodes, and a rash. Monkeypox disease is typified by rash progression from characteristic macular through papular, vesicular, and pustular phases (Damon, 2011). The duration of illness is typically 2-4 weeks and human infection has a case fatality rate of approximately 3.6% for the West African clade of MPXV and 10.6% for the Central African clade of MPXV (Durski *et al*., 2018; Beer and Rao, 2019; Bunge *et al*., 2022).

The animal reservoir for MPXV is not fully understood; however, it is likely that both large mammals and small rodents play a role in the natural zoonotic cycle (Doty *et al*., 2017; Alakunle *et al*., 2020). Primary human cases of monkeypox likely occur due to a spillover event from contact with an infected mammal (Damon, 2011). MPXV can be transmitted between people through contact with infected bodily fluids or lesions, and possibly via large droplet, aerosol, and/or fomite transmission; however, person-to-person transmission is not frequent with an estimated R_0_ of <1.0 (Alakunle *et al*., 2020).

The endemic regions for MPXV are Central and West Africa with the majority of human cases reported from the Democratic Republic of Congo since its first identification as a human pathogen in 1970 (Durski *et al*., 2018; Sklenovská and Van Ranst, 2018; Beer and Rao, 2019). In 2017, human cases of monkeypox were reported in Nigeria for the first time since the late 1970s (Yinka-Ogunleye *et al*., 2018); more than 400 cases were reported in the following three years implying a sustained focus of endemicity and a notable threat to human health in the region (Alakunle *et al*., 2020). During this period, imported cases of monkeypox were identified following travel to Nigeria on four occasions: two separate importations into the United Kingdom 2018 (Vaughan *et al*., 2018), one importation into Israel in 2018 (Cohen-Gihon *et al*., 2020), and one importation into Singapore in 2019 (Mauldin *et al*., 2020; Yong *et al*., 2020). Onward transmission was recorded in only one of these importations – during the second 2018 importation into the UK where a health worker treating the patient contracted monkeypox after changing the patient’s bedding and clothing without respiratory protection prior to initial diagnosis (Vaughan *et al*., 2020).

In early December 2019, a traveller in their 40’s sought medical assistance shortly after return to the UK following a 4-week trip to Nigeria. Symptoms began approximately two weeks before their return with a 4-day febrile illness followed by a persistent widespread pustular rash. Upon examination, the patient was apyrexial and felt well but had overt lesions covering many parts of the body including their genitals, the palms of the hands, and soles of the feet. They reported no known contact with animals, vermin or raw foods, and no travel to rural areas whilst in Nigeria. Blood EDTA, urine, throat swab, and swabs from three lesions were for tested at the Rare and Imported Pathogens Laboratory at UK Health Security Agency, Porton; all samples were positive when analysed by both pan-orthopox and MPXV-specific real-time quantitative PCR (RT-qPCR) assays confirming the diagnosis of monkeypox. Following diagnosis, the patient was transferred to a specialist infectious disease hospital for observation and treatment; this report focusses on the parallel work to establish risk of onward transmission for environments in which the patient spent prolonged periods of time since return to the UK and prior to diagnosis. The locations identified included adjoining residences occupied by the patient and their sibling in addition to a long-distance commercial bus used by the patient to travel approximately 200 miles from the airport to their residence.

## Methods

### Sampling locations

Sampling was undertaken in two adjoining single-room rented residences separated by a shared landing, located on the same floor of a 3-storey complex. The patient lived in one residence with their sibling living in the other. Also on this floor were two separate toilet facilities (one with a bath, the other with a shower) and a storage room used by both the patient and their sibling; it is believed that occupants on the floor above also used at least one of the toilet facilities. Separately, sampling was also undertaken on a bus used by the patient for a 4-hour journey to their residence after arrival into the UK (all samples from the bus were negative and will not be reported in further detail). Sampling was performed three days after the patient was admitted to hospital.

### Environmental sampling

Areas targeted for sampling were high-frequency touch/contact points within the residence. Non-porous surfaces were sampled using commercially available flocked swabs with Viral Transport Medium (Copan, USA); porous surfaces were sampled using a custom vacuum-based air sampler and gelatine filters (Sartorius, Germany). Swabbing was performed with even strokes applied both horizontally and vertically across the surface. Where possible, a 10cm × 10cm surface area was sampled in order to estimate level of contamination; however, many areas sampled were not conducive to standardised sampling such as door handles, light switches, and remote controls. For porous samples, a gelatine filter prepacked within a plastic housing was placed onto an adapter connected to a modified vacuum cleaner (Miele) via a hose. The vacuum cleaner was calibrated to run at 100 L/min. The gelatine filter attachment was then used to sample for potentially re-aerosolisable material for 5 mins per sample. Upon completion, the plastic housing containing the filter was detached and placed into a sterile bag for transport to the laboratory for processing.

### Personal-protective equipment (PPE)

Operators conducting sampling wore PPE including polyethylene coverall (Tvyek®), Tyvek® overshoes, and two layers of nitrile gloves. Overshoes and inner gloves were secured to the suit using duct tape. Respiratory protection was provided by Proflow® full hood respirators with HEPA filters. PPE was donned and doffed in a demarcated zone adjacent to the area being sampled. Doffing of PPE was performed in a set format with decontamination of gloves between each step (the outer pair of gloves being the first item removed; the inner pair being the last item removed).

### Sample processing

Environmental samples were processed inside a Class III Microbiological Safety Cabinet (MSCIII) in a containment level 3 (CL3). Swabs were gently squeezed against the tube to release media; filters were dissolved in 20mL of warmed MEM media (Gibco, USA) to provide samples for RT-qPCR.

### RT-qPCR

Original clinical samples relating to the patient and environmental samples were processed at the Rare and Imported Pathogens Laboratory (RIPL) at UK Health Security Agency, Porton. Clinical samples were screened for a range of viral and bacterial pathogens in accordance with standard clinical differential diagnosis for the patient’s symptoms; this analysis included analysis for orthopox viruses. All clinical samples were initially inactivated inside a MSCIII at CL3 laboratory using Roche MagNA Pure 96 external lysis buffer and extracted using the Roche MagNA Pure 96 platform by an external lysis protocol. Extracts were initially assessed using a pan-orthopox RT-qPCR assay (Schroeder and Nitsche, 2010) then typed using an MPXV-specific assay (Li *et al*., 2010). All assays were performed using TaqMan Fast Virus 1-step mastermix (ThermoFisher) and run on the Viia7 (ThermoFisher) platform with minor modifications to published mastermix composition and cycling conditions to conform with standardised diagnostic protocols. Environmental samples were inactivated using AVL Buffer followed by nucleic acid extraction using the Viral RNA mini kit (both Qiagen) in accordance with the manufacturer’s guidance. Environmental samples were only analysed using the pan-orthopox RT-qPCR assay.

### Viral isolation

Selected RT-qPCR positive environmental samples were cultured in Vero E6 cells inside an MSCIII cabinet within a CL3 laboratory. Briefly, environmental samples were clarified by centrifugation (13,000rpm for 3 min at 4°C) with 0.5mL supernatant used to inoculate Vero C1008 cells (BEI Resources Cat. No. NR-596, USA) in T-25cm^2^ non-vented tissue culture flasks (Corning, USA). A negative control flask was prepared following the same method with 0.5mL of MEM (Gibco, USA) used as inoculum. Inoculated flasks were incubated at 37°C without CO_2_ for 1 hour with occasional rocking. The inoculum was removed from all flasks and the cell monolayers were washed three times with sterile PBS, followed by the addition of 5mL growth medium consisting of 1 x MEM + GlutaMAX™ supplemented with 10% heat-inactivated FBS, 25mM HEPES, and 4x antibiotic-antimycotic solution (all Gibco, USA). All flasks were incubated at 37°C for 7 days and monitored for cytopathic effect (CPE) using a phase contrast inverted light microscope. Cell monolayers that did not display CPE were sub-cultured a maximum of three times, providing continuous assessment for CPE for approximately three weeks. Some flasks showed evidence of bacterial or fungal contamination despite utilisation of high concentration antibiotic-antimycotic solution; in such instances, media was exchanged to try and limit effect on viral isolation. Cells were harvested when strong CPE was evident using one freeze-thaw cycle followed by low-speed clarification at 3,000rpm for 10 mins at 4°C.

### Sequencing

In order to produce sufficient quantities of material to sequence, DNA samples were tagged, randomly primed and amplified using a modified sequence-independent amplification (Greninger *et al*., 2015; Wright *et al*., 2015; Kafetzopoulou *et al*., 2018). Amplified DNA samples were sent to the Central Sequencing Laboratory Colindale; libraries were prepared using the Nextera XT sequencing kit following the manufacturers instruction and run on an Illumina MiSeq. BWA MEM v0.7.17 was used to align reads to reference sequence MT903344.1; mapping consensus sequences were generated using in-house software QuasiBam. Consensus genome sequences and 23 representative references genomes were aligned using MAFFT v7.487. A maximum likelihood tree was constructed using MEGA 7.0.26 using the Tamura-Nei substitution model.

### Electron microscopy

Approximately 5-10μL of formalin-fixed culture supernatant (final formalin concentration >10%) was dispensed onto a formvar-filmed, carbon coated 400-mesh Transmission Electron Microscopy (TEM) grid. The sample was left to adsorb for 5 minutes. Excess suspension was removed by touching the edge of the TEM grid with moistened filter paper. Approximately 5-10μL of 2% methylamine tungstate solution was added to the grid as negative stain. After 15 seconds, excess stain solution was removed by touching the edge of the grid with moistened filter paper. The TEM grid was left to air dry prior to examination in a TEM (FEI/Philips CM100, operated at 80kV with a lanthanum hexaboride [LaB6] filament).

## Results

A total of 42 environmental samples (32 surface swabs and 10 vacuum samples) were collected from residences of the patient and their sibling including two separate single-room accommodations, two bathroom facilities, and a landing area linking these rooms. All locations were on the same floor of a multi-storey apartment complex; the only other room present on this floor was a large utility room which was not sampled as it was used for long-term storage and was not frequently accessed. Both swab samples and vacuum-based sampling methodologies were employed during this outbreak investigation to maximise the recovery efficiency from both porous materials such as bedding, sofas, and towels as well as non-porous materials such as door handles and toilet seats.

Of the 42 samples collected, 37 (88.1%) were confirmed as positive for MPXV DNA via RT-qPCR with crossing threshold (Ct) values ranging from 22.6 to 38.1 (Table). All 21 samples collected from the patient’s single-room residence were positive for MPXV DNA including 14 samples with Ct values of less than 30.0 inferring a high level of contamination. Five on the six samples collected from the sibling’s single-room accommodation were also positive for MPXV DNA; however, Ct values were much higher in positive samples ranging from 30.5 to 35.3 inferring a lower degree of contamination. MPXV DNA was also frequently identified in both bathroom facilities (5/8 samples positive with Ct values ranging from 29.9 to 33.5) and from door handles, light switches, and bannisters in the landing area (6/7 samples positives with Ct values ranging from 28.1 to 38.1). These results show widespread MPXV DNA contamination not only in the patient’s main residence, but also in locations in which they spent less time including high-touchpoint areas such as door handles and light switches.

**Table:** RT-qPCR crossing threshold (Ct) values for environmental samples collected at the patient’s residence including the adjoining residence occupied by their sibling and shared bathrooms. Results have been ordered by Ct value to allow assessment of contamination level (lower Ct value indicates greater contamination with MPXV DNA). NA = not attempted. ND = not detected. Invalid = internal extraction control outside of defined parameters; negative result from these samples cannot be considered as accurate. NB Ct values for samples collected from the bus are not shown - all samples were negative for MPXV DNA.

| Room | Item sampled | Sample method | Ct value | Viral isolation | NGS data |
| --- | --- | --- | --- | --- | --- |
| Patient's residence | Mattress and sheet | Vacuum | 22.6 | ✓ | NA |
| Patient's residence | Towel (left on top of bed) | Vacuum | 22.7 | ✓ | ✓ |
| Patient's residence | Bedding (duvet and pillows) | Vacuum | 23.7 | NA | NA |
| Patient's residence | iPad | Swab | 25.2 | ✓ | ✓ |
| Patient's residence | Door handle to landing | Swab | 25.4 | NA | NA |
| Patient's residence | Watch | Swab | 25.8 | NA | NA |
| Patient's residence | Corner taps of wash basin | Swab | 25.9 | NA | NA |
| Patient's residence | Fridge | Swab | 26.9 | NA | NA |
| Patient's residence | Passport/holder/wallet | Swab | 27.2 | NA | NA |
| Patient's residence | Small towel and cushions | Vacuum | 27.7 | NA | NA |
| Landing | Handle to patient's residence | Swab | 28.1 | ✓ | ✓ |
| Patient's residence | Kitchenette cupboard handles | Swab | 28.2 | NA | NA |
| Patient's residence | TV remote | Swab | 28.5 | NA | NA |
| Patient's residence | Large suitcase handle | Swab | 28.6 | NA | NA |
| Patient's residence | Toothbrush blue (in residence) | Swab | 28.6 | NA | NA |
| Main bathroom | Toilet flush | Swab | 29.9 | NA | NA |
| Main bathroom | Sink taps | Swab | 29.9 | ✗ | NA |
| Landing | Handle to main bathroom | Swab | 30.0 | NA | NA |
| Patient's residence | Light switch | Swab | 30.0 | NA | NA |
| Patient's residence | Toothbrush pink (in residence) | Swab | 30.5 | NA | NA |
| Sibling's residence | Duvet | Vacuum | 30.5 | ✓ | NA |
| Patient's residence | Standalone tall light switch | Swab | 30.7 | NA | NA |
| Sibling's residence | Towel | Vacuum | 31.1 | NA | NA |
| Landing | Handle to utility room | Swab | 31.3 | NA | NA |
| Patient's residence | Intercom phone to front door | Swab | 31.4 | NA | NA |
| Landing | Bannister | Swab | 31.4 | ✗ | NA |
| Patient's residence | Sofa | Vacuum | 31.6 | ✓ | NA |
| Landing | Hall Light Switch | Swab | 32.0 | ✗ | NA |
| Patient's residence | Wardrobe handles | Swab | 32.0 | NA | NA |
| Main bathroom | Toilet seat | Swab | 32.1 | NA | NA |
| Patient's residence | Electronic remote | Swab | 32.2 | NA | NA |
| Sibling's residence | Pillows | Vacuum | 32.2 | NA | NA |
| Small shower room | Sink taps | Swab | 32.5 | NA | NA |
| Main bathroom | Inner door handle | Swab | 33.5 | NA | NA |
| Sibling's residence | Mattress and sheet | Vacuum | 34.3 | ✗ | NA |
| Sibling's residence | Towel (left on top of bed) | Vacuum | 35.3 | NA | NA |
| Landing | Bedding (duvet and pillows) | Swab | 38.1 | NA | NA |
| Landing | iPad | Swab | ND | NA | NA |
| Sibling's residence | Door handle to landing | Swab | ND | NA | NA |
| Small shower room | Watch | Swab | ND | NA | NA |
| Small shower room | Corner taps of wash basin | Swab | Invalid | NA | NA |
| Small shower room | Fridge | Swab | Invalid | NA | NA |

All 10 vacuum samples collected were positive for the presence of MPXV DNA including the three samples that produced the lowest Ct values. Of the 32 swab samples collected, 27 were identified as positive for MPXV DNA. Only 5/42 samples (11.9%) returned a negative RT-qPCR result; however, two of these samples were invalid due to inhibition detected in the internal control (MS2 bacteriophage RNA spiked prior to extraction) meaning amplification of any MPXV DNA potentially present in these two samples could have been similarly inhibited.

As the presence of MPXV DNA does not necessarily infer onward transmission risk, viral isolation was attempted on a selection of RT-qPCR positive samples from different locations throughout the rooms located on this floor of the apartment complex. All four isolation attempts from samples collected in the patient’s residence were successful (Ct values for original samples ranging from 22.6 to 31.6), in addition to successful isolation from a sample collected at the sibling’s residence (original Ct value 30.5) and the external handle to the patient’s residence located on the landing area (original Ct value 28.1). CPE observed in infected cell monolayers was cell rounding, cell condensation and, ultimately, detachment. Isolation attempts were unsuccessful in four samples: two from the landing area, one from the sibling’s residence, and one from the main bathroom (original Ct values ranging from 29.9 to 34.3). Collectively, these results show the MPXV can be readily isolated from strongly positive environmental samples although the likelihood of success decreases when Ct values are greater than ∼30.

Transmission electron microscopy was performed on virus isolated from a towel in the patient’s residence (original Ct value 22.7). The resulting image (Figure 1) shows a classical ‘Mulberry’ form pox virion approximately 300nm × 200nm in size with visible surface tubules.

**Figure 1:**
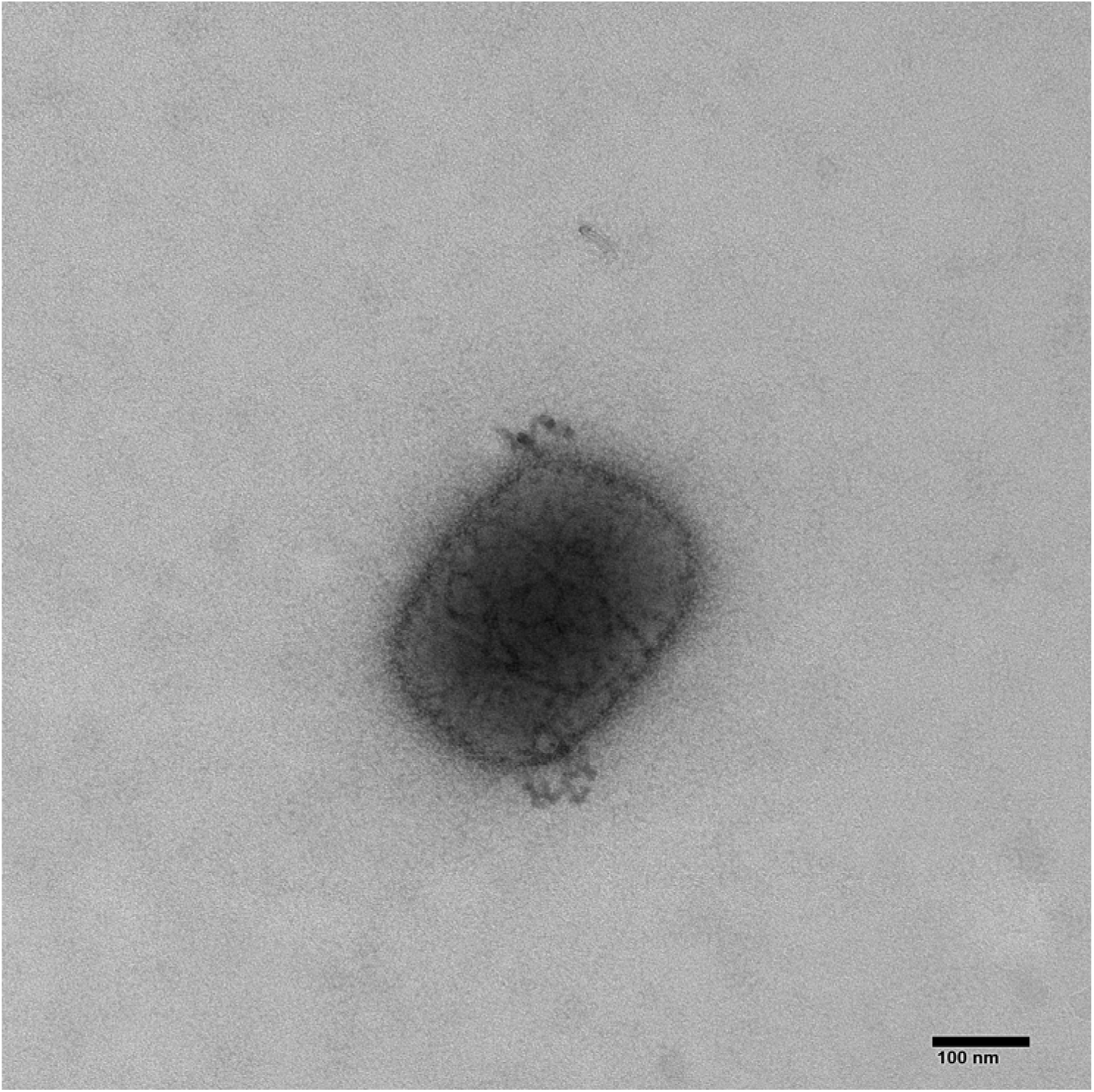
Transmission electron micrograph of Mulberry form pox virion cultured from a towel sampled in the patient’s bedroom. Direction magnificent = x92000.

Next-generation sequencing analysis was performed on viral supernatant from three samples that produced notable CPE to gain phylogenetic information regarding the provenance of the virus: a vacuum sample from towel on patient’s bed (in-house ID 2091), a swab sample from an iPad in patient’s room (in-house ID 2095), and a swab from the handle to patient’s residence (in-house ID 2102). All three samples yielded complete West African clade MPXV genome data showing close similarity to previous sequences deposited within GenBank relating to recent isolates from Nigeria including three sequences relating to previous monkeypox cases imported into the UK (Figure 2). These sequences were uploaded to Genbank with the assigned accession numbers: OL504741-OL504743.

**Figure 2:**
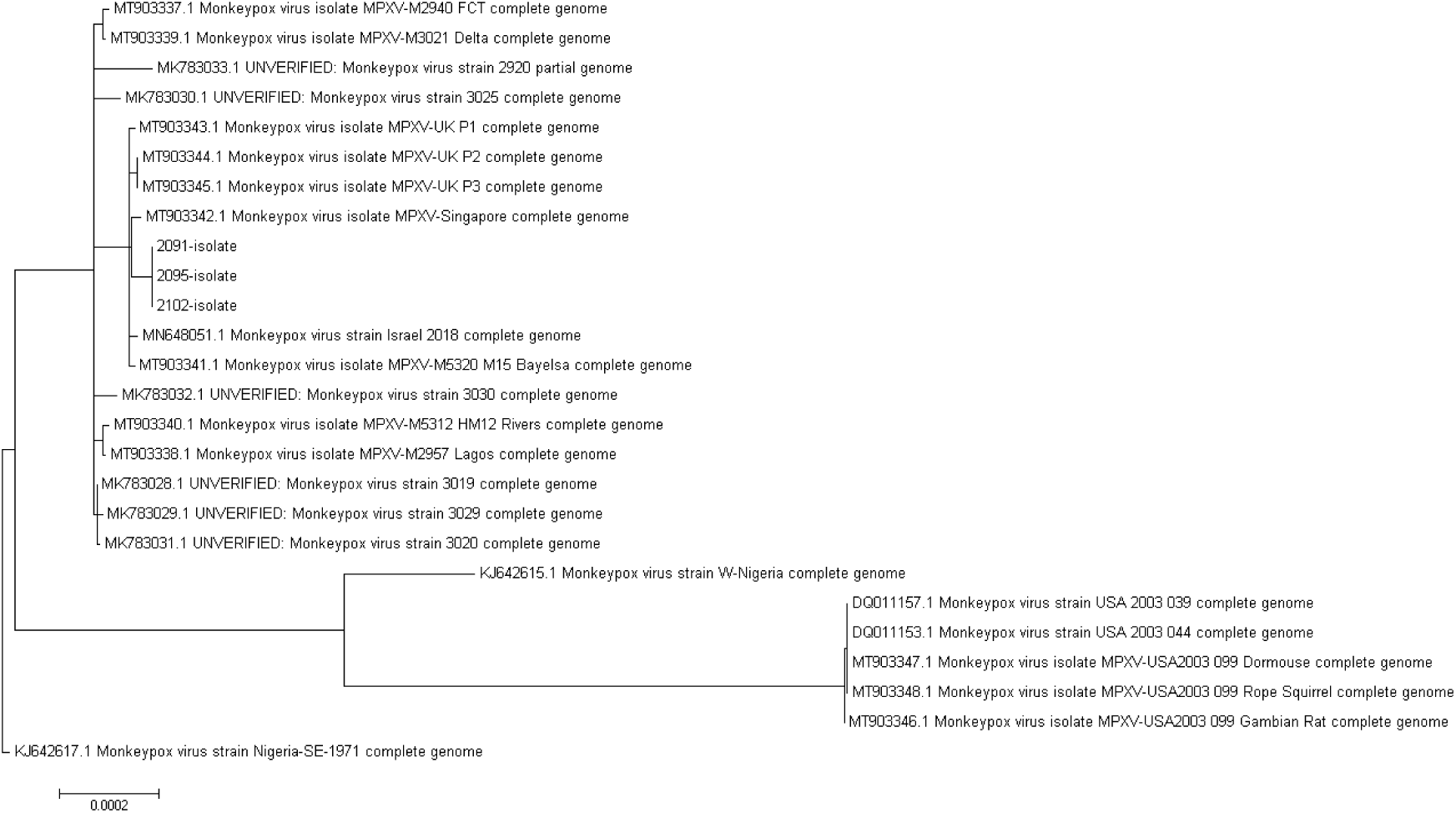
Phylogenetic tree for three environmental samples in comparison to other published complete MPXV genomes. Scale bar represents 0.0002 nucleotide substitutions per site. 2091 = vacuum sample from towel on patient’s bed (viral supernatant collected 10 days post infection when CPE developed); 2095 = swab from iPad in patient’s room (viral supernatant collected 10 days post infection when CPE developed); 2102 = swab from handle to patient’s residence (viral supernatant collected 10 days post infection when CPE developed). Sequence assigned GenBank accession numbers OL504741-OL504743.

## Discussion

In December 2019, an imported case of monkeypox was detected in a patient shortly after return from Nigeria. In addition to patient care and contact tracing, the public health response to this identification included environmental sampling both at the residence of the patient and the bus on which they travelled for several hours. Although MPXV is not considered to be easily transmitted person-to-person, effective risk analysis and contact tracing is critical to establish the risk of exposure to other family members, friends, and members of the public.

The sampling undertaken in the patient’s residence, the adjoining residence occupied by the patient’s sibling, two shared bathrooms, and the connecting landing area showed significant contamination with MPXV DNA. All 21 surface samples collected from the patient’s residence were RT-qPCR positive with most samples showing Ct values consistent with significant contamination. Numerous positive samples were also identified in sibling’s residence, shared facilities, and the connecting landing. The contamination detected in the sibling’s residence likely occurred from time the patient spent in this room playing on a games console with their sibling.

The identification of MPXV DNA from multiple environmental samples was confirmed using RT-qPCR with viral isolation used to establish whether infection-competent material was present. Monkeypox virus was successfully isolated from six samples demonstrating that infectious virus was present at the time of sampling which was three days after the patient had been admitted to hospital. The presence of viable virus could not be confirmed in four samples selected for viral isolation; all four samples had Ct values ≥29.9 which may indicate an estimate for prioritising samples for viral isolation attempts for future sampling responses; however, two of the samples yielding viable virus had Ct values greater than 29.9 indicating that this value is only an approximation. Importantly, Ct values alone cannot be used to infer infection potential and viral isolation attempts are required to ascertain presence of infectious virus in environmental samples.

Electron microscopy and whole genome sequencing was utilised to provide further confirmation and characterisation for positive samples. Classic pox-like virions were observed using transmission electron microscopy and sequence data from several successful isolations indicated that this virus was closely related to sequences associated with recent MPXV importations from Nigeria including three previous sequences from human infections from travel-related importations into the UK.

In addition to the two adjoining residences occupied by the patient and their sibling, sampling was also conducted on the public bus used by the patient following their arrival back into the UK. All samples from this bus (including the patient’s seat and surrounding seats, overhead bin, hand holds, and the onboard toilet) were negative for MPVX DNA via RT-qPCR. While this result infers the absence of possible fomite transmission following the journey taken by the patient, it is important to note that sampling of this bus was performed approximately one week after the journey was made and that the bus would have been cleaned during this time. No monkeypox symptoms were reported by passengers travelling on the bus at the same time as the patient, or those that subsequently used the bus.

The ability to sample areas potentially contaminated with high-consequence pathogens is of vital importance in order to maintain community health and public safety. This report details the response to an imported case of monkeypox into the UK from the perspective of environmental contamination and the risk of onward transmission. Despite the supposed low probability of onward transmission, effective clinical diagnosis and public health response including contact tracing is imperative as the risk is non-zero as highlighted by onward transmission to a health care professional documented in one of the previous importations. One of the potential transmission risk activities performed by the affected health care professional was changing the bedding of the patient (Vaughan *et al*., 2020); the sampling reported here identified the patient’s bedding yielded the most positive RT-qPCR signal and MPXV was isolated from this sample. While the risk of secondary transmission is believed to occur typically through prolonged close contact, it is possible that specific activities with potential to aerosolise virions can contribute including activities such as changing contaminated bedsheets, vacuum cleaning, and transport of contaminated items for washing. Utilisation of a modified vacuum sampler in this study proved effective at recovering infection competent MPXV virions from porous materials using high volumetric flow rates. Upon completion of the environmental assessment, local decontamination of the two premises was performed in accordance with current UK guidance (Public Health England, 2018).

Since this imported case was reported in December 2019, several further importations of MPXV have been identified. In May 2021, three members of the same family were diagnosed with monkeypox in the UK (the index case following travel to Nigeria, and two close family members following prolonged close contact prior to diagnosis of the index case) (Hobson *et al*., 2021). In addition, two separate importations of monkeypox into the United States were reported in 2021 following travel to Nigeria (Costello *et al*., 2022; Rao *et al*., 2022). In May 2022, an imported case of monkeypox was identified in the UK in a patient returning from Nigeria (UKHSA, 2022). Later that month, numerous cases of monkeypox were identified in the UK without travel history to MPXV-endemic regions; subsequent investigations inferred direct contact transmission with cases predominantly in gay, bisexual, and other men who have sex with men (GBMSM). Reporting of these findings led to several other countries identifying similar cases (Cohen, 2022).

In total, there have been at least nine separate importations of monkeypox following travel to Nigeria in less than four years since September 2018 (five into the UK, two into the United States, and one each into Singapore and Israel) excluding the as yet unknown origin of the 2022 cases linked predominantly in GBMSM. These cases align with re-emergence of MPVX in Nigeria likely tied with increased urbanisation and decreasing immunity in the region previously provided from smallpox eradication campaign vaccination (Rimoin *et al*., 2010; Nguyen *et al*., 2021). Given the number of clinical cases of monkeypox reported in Nigeria since 2017, combined with the number of imported cases from this region reported in the subsequent five years, it is predictable that future importations will occur with potential for onward transmission. Any country with significant transport or travel links with known endemic areas should prepare for such situations including the ability to identify and respond to clinical cases in additional to assessing the level of contamination in affected locations. Countries also need to be prepared for cases unrelated to travel to endemic regions as demonstrated by unrecognised spread of MPXV to multiple countries in 2022. This report confirms that infection-competent MPXV can be recovered from environmental settings associated with positive clinical cases that may present a risk of onward transmission to close contacts or members of the general public present in these locations. The methods utilised in this investigation for environmental sampling, RT-qPCR identification and viral isolation of positive samples can be utilised as part of the public health response activities when cases of this high consequence human pathogen are identified in future.

## Data Availability

All data produced in the present study are available upon reasonable request to the authors

## Acknowledgments

The authors wish to acknowledge the numerous clinical consultants and medical professionals from multiple locations who led the clinical response that was conducted in parallel to the work reported here. Information provided from the clinical response was paramount for informing the environmental sampling required.

The authors also wish to acknowledge assistance from Jane Osborne, Howard Tolley, Steve Pullan, the Medical Affairs group, and laboratory staff within the Rare and Imported Pathogens Laboratory for accommodating samples and requests relating to this report into their regular work.

## Declarations

### Ethical approval

The environmental sampling results presented are from an urgent public health investigation performed as part of UKHSA’s public health incident response to the importation of a high consequence infectious disease. UKHSA is the national public health agency for England and an executive agency of the UK Government’s Department of Health. The study protocol was subject to retrospective internal review by the Research Ethics and Governance Group, which is the UKHSA Research Ethics Committee; it was determined that ethical approval was not required. Information pertaining to the clinical case was provided by the Rare and Imported Pathogen Laboratory also located within UKHSA; in the context of communicable diseases, UKHSA has legal permission, provided by Regulation 3 of The Health Service (Control of Patient Information) Regulations 2002, to process confidential patient information with a view to controlling and preventing the spread of such diseases and risks, and monitoring and managing outbreaks of communicable disease.

### Authors’ contributions

Conceptualisation and methodology: BA, CB, TP, TB, AMB and KSR.

Investigation: CB, TP, K-AT, DN, AC, JP, JF, TB, AMB and KSR

Formal analysis: BA, CB, TP, JP, SS, KL, JF, KD, TB, AMB and KSR

Writing – original draft: BA, TP, JF, AMB and KSR

Writing – review and editing: All authors

